# Assessing preventive health counseling during pediatric well-child visits for children ages 6–12: A systematic review of duration and content

**DOI:** 10.1101/2025.10.15.25338080

**Authors:** Alfiya Shaikh Mohd Rafiq, Anindita Roy, Wardah Khawaja, Aishah Shaikh, Saylor Mealing, Rebecca Jungbauer, Lisa Bailey-Davis, Margret J. Foster, Kyle M. Holland, Joshua Yudkin

**Author notes:** Corresponding Author: Alfiya Shaikh Mohd Rafiq^1^, MSc, Texas A&M University, School of Public Health, 212 Adriance Lab Road, College Station, TX 77843, USA.

## Abstract

**Background:** Well-child visits (WCVs) are critical for preventive counseling but often constrained by limited time and variability in delivery. This study systematically reviews U.S.-based research published between 2014 and 2025 to examine the duration and content of preventive counseling provided during WCVs for children aged 6–12 years, with a particular focus on practitioner communication related to overweight and obesity.

**Methods:** We conducted a systematic review of U.S.-based studies that was reported in accordance with the PRISMA 2020 guidelines. Comprehensive searches were conducted in MEDLINE (Ovid), Embase (Ovid), and CENTRAL. Two reviewers independently screened titles, abstracts, and full texts, with discrepancies resolved by discussion or a third reviewer. Data was extracted using Covidence with a standardized form, and analyses were conducted in Microsoft Excel. Key variables included counseling content, duration, and delivery approach.

**Result:** After screening 2,588 references, seven studies met inclusion criteria. Preventive counseling addressed nutrition, weight management, physical activity, behavioral health, cardiovascular risk, and injury prevention, but coverage was inconsistent, with most studies reporting only a subset. Missed opportunities were common, particularly for cardiovascular risk, injury prevention, and follow-up counseling. No study quantified the duration of individual topics; however, one study reported that visits lasting ≥15–20 minutes were associated with higher odds of counseling on injury prevention (OR = 2.8), nutrition (OR = 3.0), and physical activity (OR = 6.5). Language-concordant care was limited, with interpreters used in only 25% of applicable visits. Two studies described engagement strategies such as motivational interviewing or structured follow-up, and only one reported transdisciplinary care. Notably, one large cohort study linked electronic health record (EHR) documentation of weight management counseling with improvements in child BMI, suggesting that documentation may reflect outcomes despite validity concerns.

**Conclusion:** Preventive counseling during pediatric WCV remains inconsistent, often lacking depth and Preventive counseling during pediatric WCVs remains inconsistent, often lacking depth and standardized reporting. Future research should prioritize culturally responsive approaches, team-based care models, and development of standardized metrics for counseling time and content. Addressing language barriers and integrating transdisciplinary teams are essential steps toward delivering equitable, high-quality preventive care, particularly for underserved populations.

**Trial registration:** Not applicable.

**Systematic review registration:** PROSPERO CRD42025064475.

## Background

Pediatric well-child visits (WCVs) provide structured opportunities for preventive health counseling during critical developmental periods. Approximately 86.5% of U.S. children attend at least one WCV annually,[1] emphasizing the extensive influence of these encounters. The American Academy of Pediatrics (AAP) recommends yearly WCV for children and adolescents aged 3 to 21 years to promote health literacy, screen for health conditions and developmental delays, and assess family functioning.[2] For children aged 6-12 years, these visits coincide with middle childhood, a pivotal developmental phase characterized by rapid cognitive, social, and physical changes that lay the foundation for health behaviors extending into adolescence and adulthood, shaping lifelong health behaviors.[3] These visits provide unique opportunities for counseling as children gain health literacy and independence in decision-making.[3]

Essential counseling domains include nutrition, physical activity, injury prevention, screen time, social-emotional development, and preparation for puberty,[4] with pediatricians playing a critical role in guiding lifelong healthy behaviors.[5] In practice, WCVs also include immunizations, growth monitoring, behavioral health screening, and anticipatory guidance.[2, 6]

The importance of these visits has been further emphasized by the persistent U.S. childhood obesity epidemic,[7] with 32.5% of children aged 10–17 living with overweight or obesity (OW/OB),[8] and 19.7% of children aged 2-19 classified as obese.[9] The COVID-19 pandemic further accelerated weight gain trajectories in children and adolescents.[10] These trends have significant long-term health implications including increased risk of adult obesity[11] and associated cardiovascular complications, emphasizing the critical role of WCVs in mitigating chronic disease consequences.[12, 13]

Effective preventive counseling requires adequate time allocation and comprehensive content delivery to achieve meaningful behavioral change.[14] However, research indicates significant limitations in both counseling duration and content in real-world practice. Visits often last 10–20 minutes, with fewer than one in five exceeding 20 minutes.[15] Longer visits are associated with improved care quality, including more comprehensive preventive messages and greater parental engagement.[14–16]

Similarly, preventive health counseling content also shows marked disparities in delivery frequency and duration. While nutrition, growth, and safety counseling are commonly addressed, topics such as physical activity and tobacco exposure receive less frequent and briefer attention.[14] Comprehensive analysis of WCV delivery reveals that clinicians address 7.2 health supervision and anticipatory guidance topics per visit, representing approximately 42% of Bright Futures-recommended age-specific topics.[16] Under-addressed topics include family support, parental well-being, behavior and discipline, physical activity, screen time, risk reduction and substance use, puberty and sexuality, social-peer interactions, and violence prevention.[16] Additionally, only 38.9% of visits begin with an open-ended question about parent or child concerns, despite this being a recommended practice for family-centered care.[16]

The American Academy of Pediatrics[4, 17] and the U.S. Preventive Services Task Force[18] recommend routine counseling on nutrition, physical activity, screen time, sleep hygiene, and mental health during these visits.[3, 4, 19] These encounters offer critical opportunities to promote healthy, active living and guide evidence-based parenting strategies throughout the formative middle childhood period. Quality metrics now emphasize family-centered outcomes in addition to biomedical indicators.[20]

Despite longstanding recommendations and the central role of WCVs in pediatric prevention, empirical evidence describing the frequency, duration, and quality of such counseling in real-world settings remains limited.[4, 18, 21, 22] Recent data quantifying preventive health counseling during WCVs for children aged 6 to12 years are particularly scarce, hindering efforts to evaluate current practices and develop targeted interventions. This systematic review aims to (1) characterize visit duration across healthcare settings and practitioner types, (2) identify the frequency and range of counseling topics, (3) assess associations between visit duration and counseling comprehensiveness, and (4) explore factors influencing counseling delivery. Findings will inform evidence-based recommendations to optimize preventive counseling, support clinical standards, and strengthen healthcare systems to improve child health outcomes during middle childhood.

## Methods

### Study design and registration

We conducted a systematic review of U.S.-based studies on preventive health counseling during pediatric WCVs for children aged 6–12 years. The study protocol was registered in PROSPERO on 29 January 2025 under protocol ID CRD42025064475.

### Eligibility criteria

Summarized in Table 1. Briefly, we included studies that (1) involved children aged 6–12 years, (2) examined preventive counseling delivered during WCVs in primary care, (3) provided quantitative or qualitative data on counseling content or duration, (4) were conducted in the United States, and (5) were published in English between 2014 and May 2025. Exclusion criteria included case reports, study protocols, commentaries, editorials, non-peer-reviewed studies, and studies conducted outside primary care. Studies lacking counseling content or focusing only on physical exams or medical interventions were also excluded

**Table 1.**
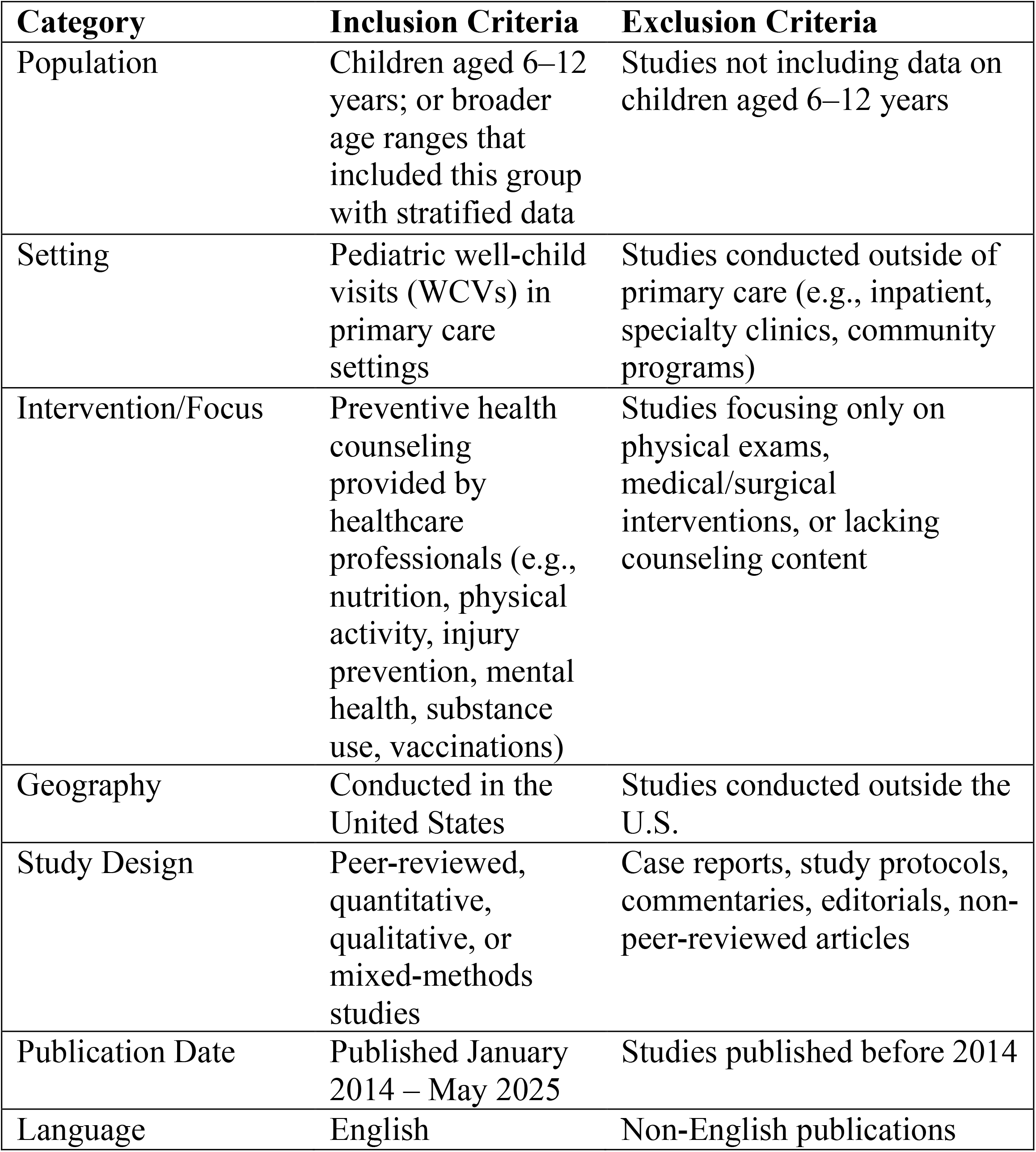
Eligibility criteria for inclusion and exclusion of studies.

| **Category** | **Inclusion Criteria** | **Exclusion Criteria** |
| --- | --- | --- |
| Population | Children aged 6–12 years; or broader age ranges that included this group with stratified data | Studies not including data on children aged 6–12 years |
| Setting | Pediatric well-child visits (WCVs) in primary care settings | Studies conducted outside of primary care (e.g., inpatient, specialty clinics, community programs) |
| Intervention/Focus | Preventive health counseling provided by healthcare professionals (e.g., nutrition, physical activity, injury prevention, mental health, substance use, vaccinations) | Studies focusing only on physical exams, medical/surgical interventions, or lacking counseling content |
| Geography | Conducted in the United States | Studies conducted outside the U.S. |
| Study Design | Peer-reviewed, quantitative, qualitative, or mixed-methods studies | Case reports, study protocols, commentaries, editorials, non-peer-reviewed articles |
| Publication Date | Published January 2014 – May 2025 | Studies published before 2014 |
| Language | English | Non-English publications |

### Search strategy

The search strategy was structured around four core concepts: persons ages six to twelve, well checks, practice patterns or patient relations, and pediatric physicians. For each concept, synonyms were identified and combined with appropriate thesaurus terms for use in each of the three databases searched: MEDLINE (Ovid), Embase (Ovid), and CENTRAL (Cochrane Library. Detailed search strategies are provided in Additional file 1. An initial search of MEDLINE (Ovid) was conducted on February 21, 2025. The retrieved references were assessed for relevance, and insights from this evaluation informed refinements to the subsequent Embase (Ovid) and CENTRAL searches.

These searches, along with an updated MEDLINE (Ovid) search, were performed on August 5, 2025. Search limits included studies published in English, within the period 2014-2025, and excluding those involving animals. All references were imported into Covidence, where duplicates were identified and removed.

### Screening and data extraction

Two reviewers independently screened titles/abstracts and full texts in Covidence, with discrepancies resolved through discussion or, when necessary, adjudication by the principal investigator (PI). To ensure consistency, the PI provided the study protocol, developed a codebook, and trained the review team. Data extraction was performed using a structured template in Additional file 2, which captured study design, health system context, provider and patient characteristics, and visit-level measures such as counseling duration, measurement method, and topics addressed. We extracted data based on sample characteristics that were stratified by practitioner and patient attributes, and study characteristics organized in terms of program format and delivery, geographic setting, study period and eligibility criteria, study funding, counseling content, and counselling duration, and counseling themes addressed. No conflicts arose during the data extraction process. Extracted data were exported from Covidence into Microsoft Excel (Microsoft Corporation, Redmond, WA, USA) for analysis and summarized for presentation in the Results section.

### Risk of bias and strength of evidence

Risk of bias for the included studies was assessed using the Joanna Briggs Institute (JBI) Critical Appraisal Checklist for Analytical Cross-Sectional Studies, given that all eligible articles employed cross-sectional or retrospective observational designs. The checklist evaluates eight domains, including clarity of inclusion criteria, description of study subjects and setting, validity and reliability of exposure and outcome measurements, identification and management of confounding factors, and appropriateness of statistical analyses. Each domain was rated as *Yes, No, Unclear*, or *Not Reported (NR)*. An overall risk of bias rating (*low, moderate*, or *high*) was then assigned to each study based on the cumulative assessment. In addition, the AHRQ Strength of Evidence (SOE) grading approach was applied to evaluate the overall quality and confidence in the body of evidence, considering study limitations, consistency, directness, and precision.

### Use of AI assistance

Large language model (LLM) tools (ChatGPT, OpenAI, 2025) were used to support language refinement and formatting of this manuscript. All authors reviewed and verified the content, and final responsibility for the manuscript rests entirely with the authors.

## Results

### Study Selection

A total of 2,588 records were initially imported for screening. 57 duplicates were removed (five identified manually and 52 via Covidence software), leaving 2,531 unique studies for review. After title/abstract screening, 2,473 records were excluded. At full-text review, 58 articles were assessed, of which 51 were excluded for the following reasons: not meeting peer-review or availability criteria (n=5), being outside pediatric primary healthcare settings or involving the wrong patient population (n=8), lacking quantitative or thematic descriptions of preventive counseling (n=16), and not meeting the specified age range of 6–12 years or lacking stratified data (n=22). A total of seven studies met all inclusion criteria and were included in the final synthesis.[20, 23–28] The study selection process is summarized in the PRISMA flow diagram (Figure 1). A detailed overview of the studies included is presented in Table 2.

**Table 2.**
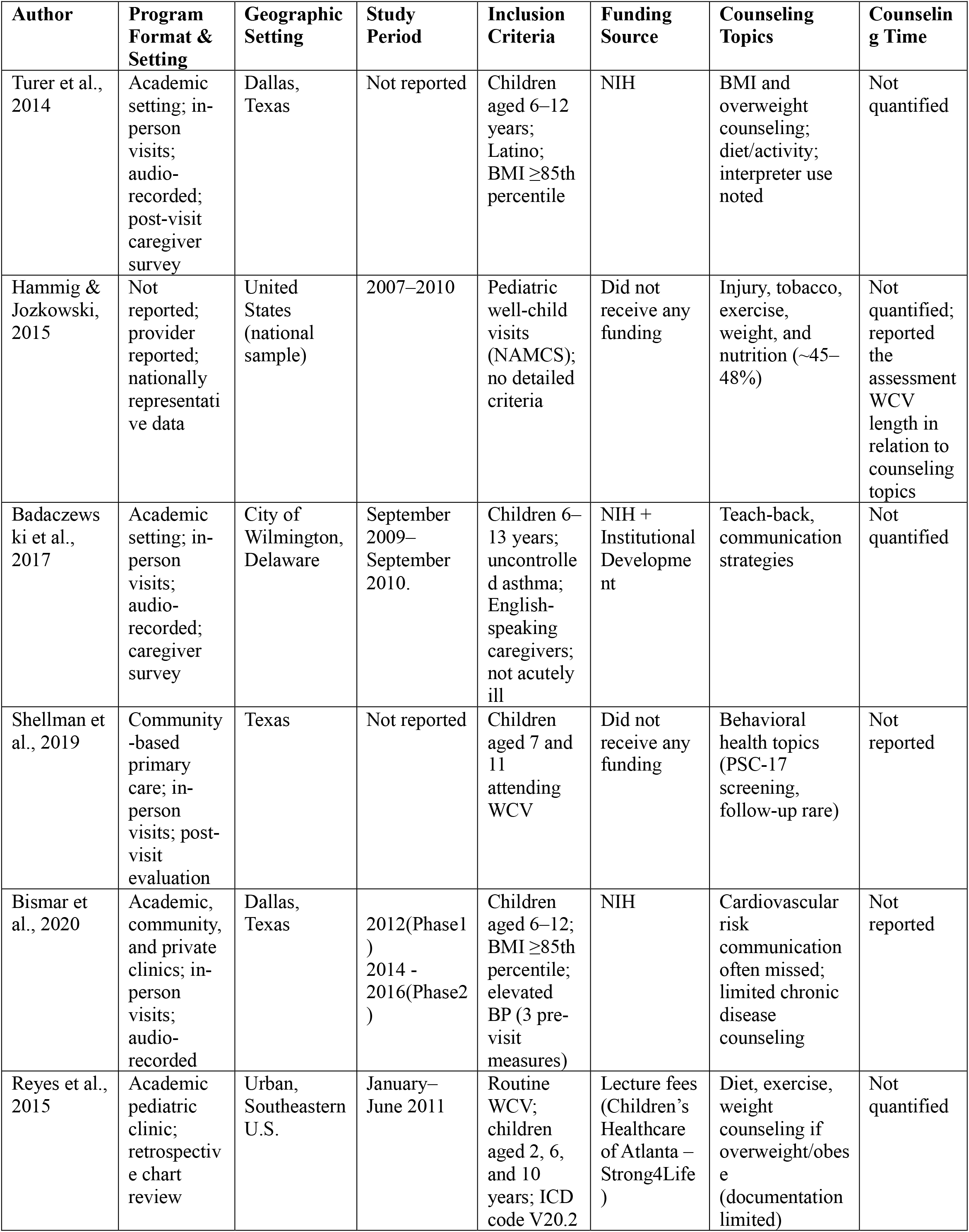
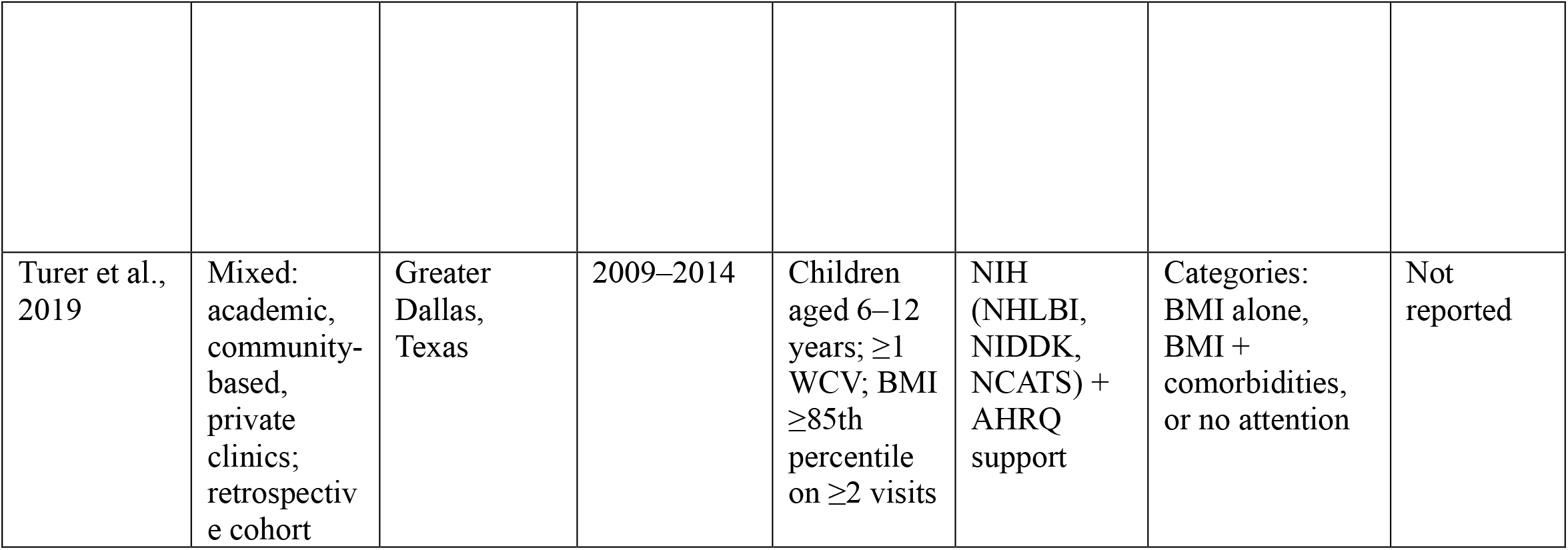
Characteristics of included studies.

**Figure 1.**
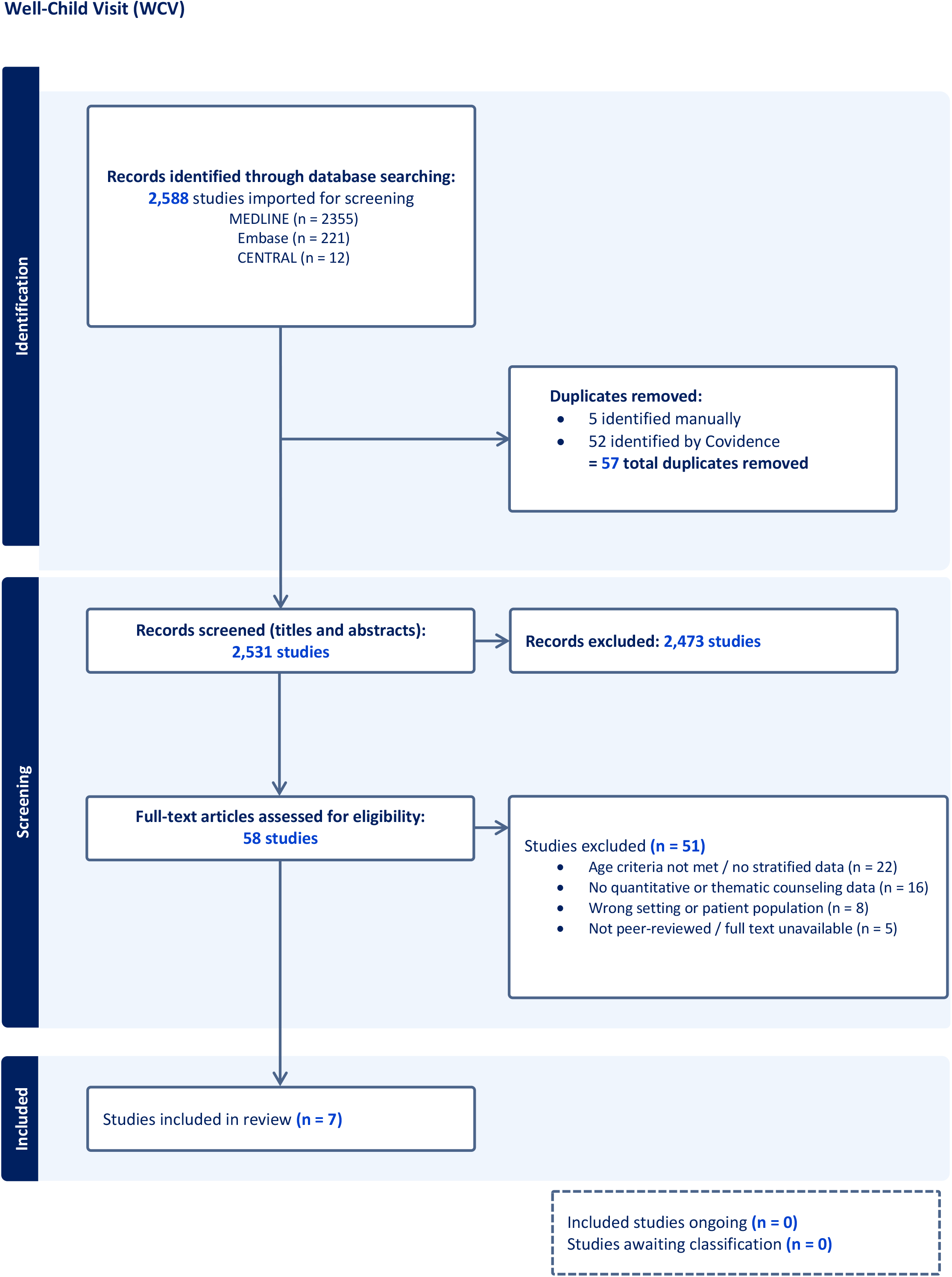
PRISMA 2020 flow diagram of study selection process.

### Socio-demographic Characteristics

#### Practitioner Characteristics

Across the seven studies, practitioner characteristics included sex (as reported in male/female terms), race/ethnicity, language of care, training background, years of experience, and self-efficacy in delivering preventive counseling. Two studies (29%) reported on practitioner gender: one study specified that 11 of the 15 pediatricians (73%) were female,[27] while another acknowledged participation by both male and female practitioners without detailing proportions.[20] Only one study reported practitioner race and ethnicity, with pediatricians self-identifying as White (40%), Asian (33%), Latino (13%), and African American (13%).[27] Two studies (29%) reported on the languages in which practitioners deliver clinical care. One study[27] found that 27% of pediatricians can deliver care in Spanish, while another[20] noted English as the sole language of delivery.

Practitioner training was described in five studies (71%), with most pediatric practitioners (hereafter “practitioners” holding MD degrees and practicing within multidisciplinary teams.[20, 23, 25–27] One study described a multidisciplinary team composed of full- and part-time pediatricians, nurse practitioners, medical assistants, and behavioral health specialists[26]. In contrast, another study only specified that practitioners were trained in pediatrics or family medicine, without further detail.[24] Two studies[20, 23] reported on years of practitioner experience: one study[20] noted that all four pediatricians had practiced for more than ten years, while the other[23] found that 18 practitioners had fewer than five years of experience and seven had more than five years. A separate study included five practitioners (four physicians and one nurse practitioner), but did not specify years of experience.[25] One study did not report any practitioner characteristics.[28] Notably, no study directly assessed practitioner self-confidence or counseling efficacy using validated measures. A summary of practitioner characteristics is presented in Table 3.

**Table 3.** Practitioner characteristics across included studies.

| Author | No. of Providers | Experience | Training & Language(s) | Sex | Race/Ethnicity | Self-Confidence / Counseling Efficacy |
| --- | --- | --- | --- | --- | --- | --- |
| Turer et al., 2014 | 15 pediatricians | Not reported | All pediatricians (MD); 27% fluent in Spanish | 73% female | White (40%), Asian (33%), Latino (13%), Black (13%) | Not assessed |
| Hammig & Jozkowski, 2015 | Not specified | Not reported | Trained in pediatrics or family medicine; language not reported | Not reported | Not reported | Not assessed |
| Badaczewski et al., 2017 | 4 pediatricians | >10 years of practice | All pediatricians (MD); English only | Male and female (not detailed) | Not reported | Not assessed |
| Shellman et al., 2019 | 16 full-time pediatricians, 4 part-time/floater pediatricians, 2 NPs, ~25 MAs, 3 psychologists, 2 trainees | Not reported | Pediatricians (MD), Psychologists, NPs; language not reported | Not reported | Not reported | Not assessed |
| Bismar et al., 2020 | 25 providers | Mixed: 18 <5 yrs, 7 >5 yrs | Pediatricians (MD); language not reported | Not reported | Not reported | Not assessed |
| Reyes et al., 2015 | 5 (4 physicians, 1 nurse practitioner). | Not reported | MD (x4), NP (x1) | Not reported | Not reported | Not reported |
| Turer et al., 2019 | Not reported | Not reported | Not reported | Not reported | Not reported | Not reported |

#### Patient Characteristics

Assessed patient characteristics included sex, race/ethnicity, parent education level, language of clinical care, insurance status, and socioeconomic status. All seven studies (100%) reported patient gender. One study[27] found a female majority (58%), five studies reported a higher proportion of male patients (ranging from 52.5% to 66%),[20, 23, 25, 26, 28] and one study reported including both genders without specifying proportions.[24] Five studies (71%) reported patient race or ethnicity. Two studies focused on Latino or Hispanic populations (one with 100% Latino[27] and another with 96% Hispanic[25]). One reported a predominantly African American sample[20], and two described racially diverse populations (one with 67% Latino[23] and 27% Black, and another with 54% Latino, 20% Black non-Latino, 14% White non-Latino, and 12% Other[28]). The remaining two studies did not report on patient race or ethnicity.[24, 26]

Parent education was reported in two studies (29%), both showing limited attainment beyond high school. One study found that 58% of parents did not complete high school[27], while another reported that among 37 respondents, 27% had less than a high school education, 32% had a high school diploma or GED, and 41% had some post-secondary education.[20] The remaining five studies did not report on parent education. Two studies (29%) reported the language in which patients received clinical care. One study facilitated visits in either English or Spanish, with 62% of families classified as limited English proficient[27] and another study reported English as the sole language of care.[20] Insurance status was reported for child participants in six studies (86%). Two studies indicated that children were publicly insured,[26, 27] while two others specifically reported Medicaid coverage.[20, 25] One study listed payer types (private, Medicare, Medicaid, and self-pay) but did not quantify the distribution,[24] and another study found that 81.1% of patients were publicly insured, 17% privately insured, and 1.9% classified as other/unknown.[28] Two studies (29%) reported parent/caregiver socioeconomic status. One study provided quantitative data, with a mean annual family income of $24,000 (95% CI: $4,500– 50,000)[27] while another study described their sample as low-income or under-resourced.[20] The remaining five studies did not report socioeconomic status. Food insecurity, a parent-reported household indicator, was not reported in any study. A summary of child and parent/caregiver characteristics is presented in Table 4.

**Table 4.** Patient and caregiver characteristics across included studies.

| Author | Sample size | Gender | Race | Parent Education | Language | Children's Insurance | Socioeconomic Status |
| --- | --- | --- | --- | --- | --- | --- | --- |
| Turer et al., 2014 | 26 children with their caregiver | Female 58% | 100% Latino | 58% < high school, 15% GED, 27% some college, 0% college graduate | English and Spanish, 62% Limited English Proficient | Public insurance | Mean annual income \$24,000 (95% CI: \$4,500–50,000) |
| Hammig & Jozkowski, 2015 | 4837 WCVs | Male and female (breakdown not provided) | Not reported | Not reported | Not reported | Private, Medicare, Medicaid, self-pay (no distribution) | Not reported |
| Badaczewski et al., 2017 | 44 children with asthma | 29 (66%) male | 98% African American | 27% < high school, 32% GED, 41% some post-secondary | English | Medicaid | Low income |
| Shellman et al., 2019 | 3678 WCVs | 51.5% male, 48.5% female | Not reported | Not reported | Not reported | Public insurance | Not reported |
| Bismar et al., 2020 | 33 children with elevated Blood pressure | Female 14/33 (42%) | Black 9/33 (27%), Latino 22/33 (67%) | Not reported | Not reported | Not reported | Not reported |
| Reyes et al., 2015 | 255 | 53.7% male (n=137), 46.3% female (n=118). | Hispanic- 96%(n=254) | Not reported | Not reported | Medicaid | Not reported |
| Turner et al., 2019 | 7,205 | Female 3,423 (47.5%)<br>Male 3,782(52.5%) | Black, non-Latino 1,454 (20.2%)<br>Latino 3,895 (54.0%)<br>White, non-Latino 992 (13.8%)<br>Other 864(12.0%) | Not reported | Not reported | Public 5,842 (81.1%)<br>Private 1,226 (17.0%)<br>Other or unknown 137(1.9%) | Not reported |

### Study characteristics

#### Program Format and Delivery Context

Six of the seven studies (86%) reported the type of primary care setting in which care was delivered. Three studies were implemented in academic settings,[20, 25, 27] two included a mix of academic, community-based, and private clinics[23, 28] and one study was conducted in community-based pediatric primary care settings.[26] Delivery modalities were primarily via in-person pediatric visits. Three studies used audio or video recordings to capture practitioner-patient communication dynamics,[20, 23, 27] while another study utilized electronic health record algorithms, validated against chart reviews and video observations.[28] In addition, three studies incorporated post-visit caregiver evaluations to assess communication quality and acceptability.[20, 26, 27]

#### Geographic Setting

All seven studies reported their geographic settings. Specifically, four studies (57%) were conducted in Texas, including two in Dallas,[23, 27] one in Greater Dallas,[28] and one in the state of Texas.[26] Additional studies were conducted in Wilmington, Delaware,[20] and in an urban clinic in the Southeastern United States,[25] while one analyzed nationally representative data from U.S. office-based practices.[24]

#### Study period and Eligibility Criteria

Study periods ranged from 2007 to 2019. Five studies reported specific years, while two did not. One study analyzed data from the 2007–2010 National Ambulatory Medical Care Survey (NAMCS),[24] another collected data between September 2009 and September 2010,[20] while a third conducted data collection in two phases: 2012 and 2014–2016.[23] One study examined a retrospective cohort from 2009 to 2014,[28] while one study reviewed charts from January to June 2011.[25] Two studies did not explicitly report the study period.[26, 27] Inclusion criteria varied across studies. Two studies enrolled children aged 6–12 years identified as overweight or obese[23, 27]; with one of these additionally requiring elevated or hypertensive blood pressure and audio-recorded visits.[23] Similarly, another study included children aged 6–12 years with BMI ≥85th percentile at two or more visits and at least one well-child visit during the study period.[28] One study enrolled children aged 2, 6, and 10 years attending routine well-child visits, though only the 6- and 10-year-old groups were retained for this review.[25] Another study restricted enrollment to children aged 7 and 11 years attending well-child visits.[26] One study recruited children aged 6–13 years with uncontrolled asthma and English-speaking caregivers, excluding those with acute illness.[20] Finally, one study included all pediatric WCVs in NAMCS without additional participant-level restrictions.[24] Taken together, three of the seven studies explicitly enrolled children with OW/OB,[23, 27, 28] and one stratified analyses by weight status, making weight-related communication a recurrent focus across the review.[25]

#### Funding Patterns

Five of the seven studies (71%) reported receiving external funding. One study was supported by the National Institutes of Health (NIH), specifically NHLBI, NIDDK, and NCATS[28] and additionally received funding from the Agency for Healthcare Research and Quality (AHRQ). Two other studies also received NIH funding.[23, 27] Another study received combined NIH and institutional development funding[20] while one study disclosed lecture fees from Children’s Healthcare of Atlanta related to Strong4Life training.[25] The remaining two studies did not report financial support.[24, 26]

### Counseling Time and Counselling Content

#### Counseling Time

Across the seven studies, none reported the average or median duration dedicated specifically to counseling. Instead, counseling time was categorized as: (1) quantified with numerical estimates, (2) described without numerical estimates, or (3) not reported. One study (14%) provided numerical estimates,[24] two studies (28%) described counseling activities without quantification,[20, 27] and four (57%) did not report counseling time at all.[23, 25, 26, 28] The only study formally analyzed visit length in relation to counseling,[24] found that longer visits (≥15–20 minutes) were associated with significantly greater odds of counseling on injury prevention (OR = 2.8), nutrition (OR = 3.0), and physical activity (OR = 6.5), but not on tobacco or weight counseling.

#### Counseling Content

The seven included studies addressed a wide range of health education domains: weight management, nutrition, physical activity, behavioral health, chronic disease risk, and preventive counseling. One study found that pediatricians discussed growth charts in 62% of visits and directly communicated overweight status in over 80% of encounters among children with BMI ≥85th percentile, often emphasizing culturally tailored nutrition and activity counseling; however, interpreter use was limited (25%).[27] Another study addressed health literacy and medication adherence, with teach-back methods used in 39% of visits.[20] A further study focused on behavioral health, conducting the Pediatric Symptom Checklist-17 (PSC-17) for psychosocial screening in 79% of visits. Follow-up counseling was rare, with additional assessments documented in less than 1.1% of cases and follow-up contacts occurring in only 1.3–2.3% of visits.[26] A nationally representative study using NAMCS data showed that counseling most often addressed nutrition (~45–48% of visits), while counseling on injury prevention, tobacco cessation, physical activity, and weight reduction was less frequent, particularly among adolescents and Black children.[24] One study identified missed opportunities for cardiovascular risk counseling, noting that in 85% of visits with overweight or obese children and elevated blood pressure, practitioners did not address cardiovascular risk.[23] Another study reported sparse counseling documentation, with only 26% of obese children and 11% of overweight children having nutrition, physical activity, or weight counseling recorded.[25] Finally, one study used electronic health record algorithms to categorize counseling as attention to BMI alone, BMI plus comorbidities, or no attention, but did not report specific content details.[28]

### Risk of bias and strength of evidence

Risk of bias was assessed using the JBI Critical Appraisal Checklist for Analytical Cross-Sectional Studies. Across the seven included studies, two were rated low risk of bias,[20, 27] four were rated moderate risk,[23, 25, 26, 28] and one was low–moderate risk due to database-related limitations.[24] Studies rated as low risk employed validated outcome measures (e.g., RIAS coding, CDC BMI cutoffs, PSC-17 behavioral screening), incorporated direct observation of counseling practices, and reported high inter-rater reliability. In contrast, moderate-risk studies were characterized by reliance on secondary data sources or documentation rather than direct observation, lack of multivariable adjustment for potential confounders, or restricted generalizability due to narrow, single-site, or demographically homogeneous populations. For example, one study did not account for pre-existing behavioral diagnoses,[26] another did not adjust for sociodemographic confounders,[23] a third risked misclassification bias by equating documentation with actual counseling.[28] and a fourth did not address practitioner- or system-level confounding.[25] A detailed, study-level summary is presented in Table 5.

**Table 5.** Risk of bias. Risk of bias was assessed using the Joanna Briggs Institute (JBI) Critical Appraisal Checklist for Analytical Cross-Sectional Studies. Each study was evaluated across eight domains: (1) clearly defined inclusion criteria, (2) detailed description of study subjects and setting, (3) valid and reliable measurement of exposure, (4) use of objective standard criteria for measurement of the condition, (5) identification of confounding factors, (6) strategies to address confounders, (7) valid and reliable measurement of outcomes, and (8) use of appropriate statistical analysis. Responses were coded as es, No, Unclear, or Not Reported (NR), with narrative justifications documented in the table. Overall risk of bias was then rated as Low, Moderate, or High based on the body of evidence and consistency cross domains.

| Author, Year | Inclusion Criteria Defined | Subjects & Setting Described | Exposure Measured Validly | Standard Criteria Used | Confounding Factors Identified | Strategies for Confounding | Outcomes Measured Validly | Statistical Analysis Appropriate | Risk of Bias |
| --- | --- | --- | --- | --- | --- | --- | --- | --- | --- |
| Turer et al., 2014 | Yes | Yes (study period not reported) | Yes | Yes | Yes (language concordance, SES, parental education, provider characteristics) | NR – stratified by language only, no multivariable regression adjusting for all confounders | Yes (validated coding, interrater reliability) | Yes ($\chi^2$ tests, kappa agreement) | Low |
| Hammig & Jozkowski, 2015 | Yes | Yes | Unclear | Yes | Yes (database lacked provider education/triage info) | Partial – limited by database | Yes (binary coding of health education delivery) | Yes | Low-Moderate |
| Badaczewski et al., 2017 | Yes | Yes | Yes (RIAS coding, validated) | Yes (CDC BMI cutoffs, AAP criteria) | Yes (language, SES, cultural factors) | Yes (language concordance, regression adjustment) | Yes (validated RIAS, Loop Score, NVS) | Yes (descriptive stats, regression, ORs/CIs) | Low |
| Shellman et al., 2019 | Yes (exclusion not detailed) | Yes (study period not reported) | Yes (PSC-17, validated psychometric tool; EMR) | Yes (PSC-17 cutoffs) | No/Unclear (behavioral diagnoses noted but not analyzed) | No (descriptive only) | Yes | Yes | Moderate |
| Bismar et al., 2020 | Yes | Yes | Yes | Yes | Yes | No/Unclear (no formal adjustment for confounders) | Yes | Yes (Fishers exact, no multivariable) | Moderate |
| Reyes et al., 2015 | Yes | Yes | Yes | Yes | No | No | Yes (clear coding rules) | Yes ($\chi^2$ tests) | Moderate |
| Turer et al., 2019 | Yes | Yes | Yes (EHR algorithm, risk of misclassification) | Yes | Yes | Yes | Yes | Yes | Moderate |

#### Strength of Evidence

The strength of evidence (SOE) was graded according to the AHRQ framework across the seven included studies.[20, 23, 25–28] Consistency was low for counseling duration, as only one study reported indirect associations between longer visits and counseling delivery, while the remaining six did not quantify counseling time. In contrast, counseling content showed moderate consistency: across multiple studies, nutrition and weight were routinely addressed, but counseling on physical activity, behavioral health, and cardiovascular risk was less frequent. Directness was moderate to high, because all studies were conducted in U.S. primary care or well-child visit settings. However, generalizability was limited because four studies were conducted in single-site academic or community clinics serving narrow populations (e.g., majority Latino or Medicaid-insured),[23, 25–27] while one study provided nationally representative estimates.[24] Precision was low to moderate: six studies were small, descriptive, and single-site (sample sizes ranging from 33 to approximately 200), with only NAMCS-based data offering stable national estimates.[24] Wide confidence intervals and the lack of repeated measures across multiple sites further limited precision.

As detailed in the risk of bias assessment, studies ranged from low to moderate risk. Equity-related findings, including disparities linked to language concordance, interpreter use, and documentation gaps among low-income and Medicaid-insured populations, were observed across three studies,[23, 26, 27] supporting moderate but limited SOE for equity considerations. Publication bias could not be excluded given the small evidence base (n=7) and the absence of trial registration or inclusion of grey literature.

## Discussion

This systematic review synthesized data from U.S.-based research over the past ten years to examine the duration and content of preventive counseling delivered during WCVs for children aged 6–12 years. Across the seven included studies, none explicitly quantified the average or median time spent on individual counseling domains.[20, 23–28] Instead, studies primarily described counseling content or documentation patterns, highlighting a critical measurement gap that impedes evaluation of counseling quality or effectiveness. Only one study indirectly linked visit duration with counseling, showing that longer visits (≥15–20 minutes) increased the likelihood of counseling on injury prevention, nutrition, and physical activity.[24] In addition, another study advanced this field by examining EHR-documented counseling behaviors and demonstrating that documentation of weight management services was associated with improvements in child BMI, though reliance on abstraction algorithms raises concerns about validity since documentation may not reflect counseling quality or depth.[28] This lack of quantification parallels residency-level evidence: fewer than half of pediatric and family medicine residents have received training in standardized developmental screening tools,[29] and while attitudes toward literacy promotion are positive, counseling practices lagged behind,[30] reinforcing the persistence of training–practice gaps. These findings align with the overall SOE rating of low strength for counseling duration and moderate strength for counseling content, emphasizing both the scarcity of precise time data and the relative consistency of nutrition-focused counseling across studies.

The limited body of research suggests significant variation and notable gaps in the consistency, depth, and delivery of preventive health counseling during pediatric visits. Across studies, most counseling was delivered in an episodic and non-standardized fashion, often initiated in response to immediate clinical findings or parental concerns rather than guided by a structured protocol, contributing to a restricted scope of counseling, with insufficient emphasis on key risk factors such as physical activity, behavioral health, and cardiovascular risk—even among children with known comorbidities like obesity or elevated blood pressure.[20, 23] Reyes et al. further highlighted this issue, reporting that only one-third of children living with overweight/obesity (OW/OB) were identified in medical records, and counseling was documented in just 11% of overweight and 26% of obese cases.[25] However, if only looking at international classification of disease (ICD) codes, OW/OB is often not be documented,[31] as some practitioners are hesitant to formally code pediatric OW/OB with an ICD diagnosis, potentially to avoid discomfort and upsetting patients.[32, 33] While data illustrate steady improvement in documenting weight assessment and counseling for children, there is still a persistent gap between actual practice and documentation, complicating accountability and quality measurement. Furthermore, unlike in primary care for adults, well-child visit reimbursement does not increase when OW/OB is documented. In practice, HEDIS quality measures remain the primary system-level mechanism supporting weight assessment and counseling.[34]

Importantly, this gap reflects a broader disconnect between practitioner practices and family needs: caregivers consistently identify mental health, nutrition, physical activity, and communication as top priorities, yet these were the least consistently addressed domains in the included studies.[35, 36] Given the moderate risk of bias in several EMR-based studies, it is likely that counseling frequency was underestimated, suggesting the real gap between practice and family needs may be somewhat smaller than reported, though still substantial.

Nutrition and weight status were the most frequently addressed topics, typically prompted by growth charts or BMI classifications. However, deeper engagement strategies—such as motivational interviewing, behavioral goal-setting, or structured follow-up—were rarely observed. Behavioral health screenings, when conducted, infrequently led to meaningful counseling or referral. This aligns with qualitative findings from a previous study, which reported that although pediatricians are confident in identifying obesity, they often lack effective strategies and referral mechanisms to manage it, resulting in limited counseling depth and outcomes.[37] Language concordance emerged as a critical influence on counseling quality. In one study, trained interpreters were used in only 25% of visits requiring interpretation, undermining effective communication and culturally-responsive care delivery.[27]

The application of health literacy sensitive strategies, such as teach-back, remains limited in pediatric practice. In one of the included studies, teach-back was used in 39% of asthma-related visits but showed no clear training benefit and was associated with negative caregiver reactions.[20] Complementary findings from international research revealed that pediatricians, though aware of the need to gauge caregiver understanding, lacked formal health literacy training and perceived teach-back as impractical due to time constraints.[38] Similarly, previous study Busch et al. demonstrated that while a brief educational intervention increased referrals and screening rates, it did not improve documentation, billing, or counseling frequency.[39] Practitioners continued to cite time pressures and parental perceptions as major barriers, reinforcing that practitioner education alone is insufficient without system-level supports.[24, 39] Evidence from other research underscores the importance of embedding structural solutions such as EHR prompts, workflow redesign, and reimbursement incentives to sustain counseling practices.[2, 40–42] Together, these findings point to the need for streamlined, evidence-based communication training that balances effectiveness with clinical feasibility.

Practitioner characteristics (e.g., gender, race/ethnicity, language skills) were inconsistently reported, limiting insight into their impact on counseling quality. Transdisciplinary teams were also underutilized, with only one study explicitly describing the involvement of behavioral health clinicians or nurse practitioners.[26] Methodologically, six of the seven studies relied on objective data sources such as audio recordings, chart reviews, or EMR abstraction[20, 23, 25–28] while only one study relied on self-reported practitioner data.[24] This raises concerns about reporting bias, given known discrepancies between reported and observed practitioner behaviors. This reflects a directness limitation identified in the SOE assessment: while all studies were conducted in U.S. pediatric settings, most were single-site, demographically narrow (e.g., Latino or Medicaid-insured populations), limiting the generalizability of findings to wider practice.

Our findings align with prior studies showing that while pediatricians may initiate discussions about weight, they often do not provide detailed counseling due to time constraints and limited training.[40] This review extends these findings by highlighting two persistent issues: the absence of data on counseling duration, which limits evaluation of intervention feasibility, and the reliance on documentation rather than direct observation, which may misrepresent counseling depth. Notably, Turer et al.’s study represents a recent contribution by linking counseling documentation to BMI outcomes and showing that preventive counseling occurs not only during well-child visits but also in non–well-child encounters.[28] It also draws attention to the historical decline in research output in this area—only one study has been published since 2020, compared to more active research over a decade ago.[23] This lack of recent literature may reflect pandemic-related disruptions, delays in dissemination, or waning academic focus despite persistent need. Publication bias cannot be excluded, given the very small number of eligible studies (n=7) and the absence of trial registration, further limiting confidence in the evidence base.

### Health Equity and Cultural Considerations

This review identified disparities in counseling delivery linked to language, race/ethnicity, and socioeconomic status. For example, one study found that interpreted visits were associated with less frequent communication about overweight status, and only 25% of such visits included trained interpreters.[27] Moreover, three studies primarily served low-income or publicly insured populations, yet few interventions were explicitly tailored to address social determinants of health such as food insecurity, caregiver literacy, or cultural dietary practices.[24, 26, 27]

Reyes et al. reinforced these equity concerns by demonstrating that under-identification and sparse documentation disproportionately affected a majority-Hispanic, Medicaid-insured clinic. The clinic’s later adoption of motivational interviewing and quality improvement (QI) initiatives illustrates that system-level changes can improve care delivery.[25] This gap in the included studies contrasts with recent external evidence: Hunt et al. demonstrated the feasibility of embedding social determinants of health screening into pediatric visits,[43] while Lui et al. piloted ADHD screening for children and parents in Medicaid-based clinics, both highlighting the importance of culturally sensitive and equity-focused interventions.[44] These findings underscore the critical need for culturally responsive and linguistically accessible counseling strategies, especially given the high burden of obesity-related disparities among Latino and African American children. In line with this, Shah et al. showed that caregivers valued culturally relevant education and trust-building in ecology-focused group visits, reinforcing the importance of tailoring WCV content to family and community context.

### Implications for Practice

Efforts to improve the quality of pediatric WCVs must consider both structural and behavioral strategies. External studies show that non-face-to-face tasks (e.g., EHR documentation, follow-up calls, and care coordination) often consume more time than direct patient interactions, thereby constraining opportunities for preventive counseling.[42] This reinforces that structural supports are critically needed to protect counseling time. Structurally, integrating electronic health record prompts, adjusting reimbursement incentives, and extending visit durations could promote more consistent and comprehensive counseling delivery. Behaviorally, enhanced practitioner training in evidence-based communication strategies such as teach-back[40] and motivational interviewing[41] grounded in cultural humility and trauma-informed care[45, 46] and tailored to cultural and linguistic diversity, may improve counseling effectiveness. Moreover, embedding transdisciplinary teams[47] including behavioral health specialists, dietitians, and health coaches— within primary care settings may help alleviate time constraints and ensure more holistic care delivery[48]. Innovative care delivery models like Centering Parenting, highlighted in external studies, demonstrate that group WCVs can reduce parental isolation, build caregiver confidence, and expand opportunities for preventive counseling, offering a scalable complement to traditional individual visits.[49]

### Implications for Policy

Given that none of the included studies explicitly reported counseling durations, there is an urgent need to develop standardized metrics and methodologies for tracking counseling time across different clinical settings. Without standardized metrics, efforts to benchmark, evaluate, or improve preventive counseling quality will remain limited. Additionally, reforms should consider revising documentation and reimbursement structures to incentivize the delivery of evidence-based preventive counseling. Policy-level reforms should build on existing systems such as NCQA’s WCC and HEDIS measures, which track BMI percentile documentation, nutrition counseling, and physical activity counseling.[34, 50, 51] Annual updates through CMS electronic Clinical Quality Measures have introduced age-stratified reporting for children and adolescents.[52] However, these measures primarily assess *whether counseling occurred* and do not capture counseling **duration, depth, or tailoring**. Future reforms should address these gaps to ensure reimbursement and documentation structures incentivize evidence-based preventive counseling. Additionally, policies that promote team-based care models and support interpreter services are critical for promoting equitable preventive counseling.

### Strengths

This systematic review has several notable strengths. To our knowledge, this is the first review to comprehensively synthesize empirical evidence on the duration and content of counseling during pediatric WCVs in the United States, specifically focusing on children aged 6–12 years—an age group and care component that have received limited attention in prior research. The review followed rigorous and transparent methods aligned with PRISMA guidelines, including dual independent screening, clearly defined eligibility criteria, and inclusion of studies employing diverse data collection methods. These methodological choices enhance the reliability, reproducibility, and generalizability of the findings while minimizing the risk of selection bias. Importantly, this review also highlights critical equity and economic considerations—such as limited use of interpreters and language barriers—that can contribute to disparities in care and increased healthcare costs. Addressing these issues is essential for promoting more equitable and efficient pediatric preventive services.

### Limitations

This review has several limitations. First, substantial heterogeneity in study designs, populations, and outcome measures limited the feasibility of conducting a meta-analysis or producing a robust quantitative synthesis. Second, many studies relied on secondary data sources or practitioner self-report, which may have led to overestimations of counseling frequency and quality—particularly in studies using data from the NAMCS, where documentation may not reflect actual practice. Third, three studies included broader pediatric age ranges without stratified results, potentially reducing the precision of findings for this target group. Lastly, there is a notable absence of studies published after 2020. This gap—potentially influenced by the COVID-19 pandemic—limits our understanding of more recent shifts in clinical practice and counseling approaches in a rapidly evolving healthcare context.

### Future Research Directions

Future research should incorporate precise time-tracking methods combined with content analysis to quantify counseling duration and quality. Validated tools measuring practitioner self-efficacy, confidence, and cultural competence should be integrated to better understand determinants of counseling delivery. Mixed methods designs, including qualitative assessments of family experiences, can enrich interpretation. Longitudinal studies exploring how practitioner training and experience influence counseling over time are also needed. Finally, intervention research must focus on scalable, culturally tailored strategies to enhance equitable preventive counseling, particularly for underserved populations with limited English proficiency.

## Conclusion

This systematic review demonstrates that preventive counseling during pediatric well-child visits for children aged 6–12 years remains limited in both duration and content. Nutrition and weight management were addressed more frequently than other topics; however, critical areas such as physical activity, behavioral health, and cardiovascular risk received inconsistent attention. None of the studies included systematically quantified counseling time, and documentation often underestimated counseling activities, limiting assessments of quality and feasibility. These findings underscore the need for standardized approaches to measuring counseling duration and content, as well as strategies to improve comprehensiveness, cultural responsiveness, and equity in preventive care. Strengthening these areas is essential to improve child health outcomes and reduce disparities during this formative developmental stage.

## Data Availability

This is a systematic review and all data are already published.

## List of Abbreviations

AAP: American Academy of Pediatrics
BMI: Body Mass Index
CDC: Centers for Disease Control and Prevention
CI: Confidence Interval
EHR: Electronic Health Record
HEDIS: Healthcare Effectiveness Data and Information Set
ICD: International Classification of Diseases
JBI: Joanna Briggs Institute
NAMCS: National Ambulatory Medical Care Survey
NCATS: National Center for Advancing Translational Sciences
NHLBI: National Heart, Lung, and Blood Institute
NIDDK: National Institute of Diabetes and Digestive and Kidney Diseases
OR: Odds Ratio
OW/OB: Overweight/Obesity
PI: Principal Investigator
PRISMA: Preferred Reporting Items for Systematic Reviews and Meta-Analyses
QI: Quality Improvement
SOE: Strength of Evidence
U.S: United States
WCV: Well-Child Visit

## Declarations

### Ethics approval and consent to participate

Not applicable (no human subjects directly studied).

### Consent for publication

Not applicable.

### Availability of data and materials

All data generated or analyzed during this study are included in this article and its supplementary information files.

### Authors’ contributions

- Alfiya Shaikh Mohd Rafiq: Led data extraction and analysis, participated in article review, and drafted much of the manuscript.
- Anindita Roy, Wardah Khawaja, Aishah Shaikh, Saylor Mealing: Participated in article review and reviewed the final manuscript.
- Rebecca Jungbauer: Led the risk of bias assessment and reviewed the manuscript.
- Lisa Bailey-Davis: Provided manuscript edits and guided contextualization of findings within the literature.
- Margret J. Foster, Kyle M. Holland: Led the search strategy and contributed to Methods section development.
- Joshua Yudkin: Conceived and conceptualized the study, oversaw its execution, and contributed to manuscript drafting and editing.

*All authors read and approved the final manuscript*

All authors have read and approved the final manuscript and agree to be accountable for all aspects of the work.

## Acknowledgements

“Not applicable”

## Additional file legends

Additional file 1. Full database search strategies (Ovid MEDLINE, Embase, CENTRAL).

Additional file 2. Data extraction template used in Covidence.

## Additional file 1. Search Strategies

### MEDLINE (Ovid) Search

#### Ovid MEDLINE(R) ALL <1946 to February 20, 2025>

1. exp Child/ or Puberty/ or ((age? adj2 (“6” or “7” or “8” or “9” or “10” or “11” or “12” or six or seven or eight or nine or ten or eleven or twelve)) or ((“6” or “7” or “8” or “9” or “10” or “11” or “12” or six or seven or eight or nine or ten or eleven or twelve) adj2 (“year? of age” or “yr of age” or “ys of age” or “yrs of age” or “y of age” or “year? old” or “yr old” or “ys old” or “yrs old” or “y old”)) or child* or kid or kids or girl or girls or girlhood or boy or boys or boyhood or juvenil* or prepube* or pubert* or pubescent? or pubescence or preadolesc* or youth? or (young adj2 (people* or person* or individual* or population*)) or schoolage? or “school age?” or schoolchild* or “school child*” or “1st grade*” or “first grade*” or “grade? 1” or “grade? one” or “2nd grade*” or “second grade*” or “grade? 2” or “grade? two” or “3rd grade*” or “third grade*” or “grade? 3” or “grade? three” or “4th grade*” or “fourth grade*” or “grade? 4” or “grade? four” or “5th grade*” or “fifth grade*” or “grade? 5” or “grade? five” or “6th grade*” or “sixth grade*” or “grade? 6” or “grade? six” or “7th grade*” or “seventh grade*” or “grade? 7” or “grade? seven” or school* or “pre school*” or “elementary school*” or “primary school*” or “postprimary school*” or “grade school*” or “middle school*” or “junior high” or “intermedia* school*” or “presecondary school*” or preschool* or gradeschool* or middleschool* or kin#ergar#en* or “elementary education*” or “primary education*” or “postprimary education*” or “intermedia* education*” or “presecondary education*” or “secondary education*” or “k through 12” or “k through twelve” or “k thru 12” or “k thru twelve” or “k 12” or k12 or “k twelve” or p?ediat*).ti,ab,kf.
2. exp Preventive Health Services/ or Preventive Medicine/ or pc.fs. or (checkup? or “check up?” or “well check?” or “wellcheck?” or ((“well child” or “health supervision” or “primary care”) and visit*) or (preven??tive adj (medicine or health or care or service? or program?)) or “health education” or “health promotion”).ti,ab,kf.
3. Practice Patterns, Physicians’/ or Physician-Patient Relations/ or Clinical Competence/ or (“practice pattern?” or “prescri* pattern?” or “patient relation*” or “physician patient” or “patient physician” or “doctor patient” or “patient doctor” or content or ((physician? or doctor? or clinician?) adj behavior?)).ti,ab,kf. or ((exp Communication/ or communicat*.ti,ab,kf.) and (Patients/ or (patient? or inpatient? or outpatient? or client?).ti,ab,kf.))
4. Physicians, Primary Care/ or Primary Health Care/ or (p?ediatrician? or (p?ediatric? adj (clinic? or practice? or setting? or hospital? or physician? or doctor? or clinician? or practice? or health))).ti,ab,kf,hw,in. or (children?? adj3 hospital?).ti,ab,kf,in. or ((“primary care” adj (provider? or physician? or doctor? or clinician? or clinic?)) or (family adj (physician? or doctor? or clinician? or clinic?))).ti,ab,kf.
5. (exp Animals/ not (exp Animals/ and Humans/)) or exp Animal Experimentation/ or exp Models, Animal/ or ((animal? adj2 (study or model* or experiment*)) or mouse or mice or rat or rats or monkey or monkeys or “preclinical study”).ti.
6. (1 and 2 and 3 and 4) not 5
7. limit 6 to (english language and yr=“2014-Current”)

### MEDLINE (Ovid) Search Update

#### Ovid MEDLINE(R) ALL <1946 to August 04, 2025>

1. exp Child/ or Puberty/ or ((age? adj2 (“6” or “7” or “8” or “9” or “10” or “11” or “12” or six or seven or eight or nine or ten or eleven or twelve)) or ((“6” or “7” or “8” or “9” or “10” or “11” or “12” or six or seven or eight or nine or ten or eleven or twelve) adj2 (“year? of age” or “yr of age” or “ys of age” or “yrs of age” or “y of age” or “year? old” or “yr old” or “ys old” or “yrs old” or “y old”)) or child* or kid or kids or girl or girls or girlhood or boy or boys or boyhood or juvenil* or prepube* or pubert* or pubescent? or pubescence or preadolesc* or youth? or (young adj2 (people* or person* or individual* or population*)) or schoolage? or “school age?” or schoolchild* or “school child*” or “1st grade*” or “first grade*” or “grade? 1” or “grade? one” or “2nd grade*” or “second grade*” or “grade? 2” or “grade? two” or “3rd grade*” or “third grade*” or “grade? 3” or “grade? three” or “4th grade*” or “fourth grade*” or “grade? 4” or “grade? four” or “5th grade*” or “fifth grade*” or “grade? 5” or “grade? five” or “6th grade*” or “sixth grade*” or “grade? 6” or “grade? six” or “7th grade*” or “seventh grade*” or “grade? 7” or “grade? seven” or school* or “pre school*” or “elementary school*” or “primary school*” or “postprimary school*” or “grade school*” or “middle school*” or “junior high” or “intermedia* school*” or “presecondary school*” or preschool* or gradeschool* or middleschool* or kin#ergar#en* or “elementary education*” or “primary education*” or “postprimary education*” or “intermedia* education*” or “presecondary education*” or “secondary education*” or “k through 12” or “k through twelve” or “k thru 12” or “k thru twelve” or “k 12” or k12 or “k twelve” or p?ediat*).ti,ab,kf.
2. exp Preventive Health Services/ or Preventive Medicine/ or pc.fs. or (checkup? or “check up?” or “well check?” or “wellcheck?” or ((“well child” or “health supervision” or “primary care”) and visit*) or (preven??tive adj (medicine or health or care or service? or program?)) or “health education” or “health promotion”).ti,ab,kf.
3. Practice Patterns, Physicians’/ or Physician-Patient Relations/ or Clinical Competence/ or (“practice pattern?” or “prescri* pattern?” or “patient relation*” or “physician patient” or “patient physician” or “doctor patient” or “patient doctor” or content or ((physician? or doctor? or clinician?) adj behavior?)).ti,ab,kf. or ((exp Communication/ or communicat*.ti,ab,kf.) and (Patients/ or (patient? or inpatient? or outpatient? or client?).ti,ab,kf.))
4. Physicians, Primary Care/ or Primary Health Care/ or (p?ediatrician? or (p?ediatric? adj (clinic? or practice? or setting? or hospital? or physician? or doctor? or clinician? or practice? or health))).ti,ab,kf,hw,in. or (children?? adj3 hospital?).ti,ab,kf,in. or ((“primary care” adj (provider? or physician? or doctor? or clinician? or clinic?)) or (family adj (physician? or doctor? or clinician? or clinic?))).ti,ab,kf.
5. (exp Animals/ not (exp Animals/ and Humans/)) or exp Animal Experimentation/ or exp Models, Animal/ or ((animal? adj2 (study or model* or experiment*)) or mouse or mice or rat or rats or monkey or monkeys or “preclinical study”).ti.
6. (1 and 2 and 3 and 4) not 5
7. limit 6 to (english language and yr=“2014-Current”)

### Embase (Ovid) Search

#### Embase <1980 to 2025 August 01>

1. (exp child/ not exp infant/) or childhood/ or ((age? adj2 (“6” or “7” or “8” or “9” or “10” or “11” or “12” or six or seven or eight or nine or ten or eleven or twelve)) or ((“6” or “7” or “8” or “9” or “10” or “11” or “12” or six or seven or eight or nine or ten or eleven or twelve) adj2 (“year? of age” or “yr of age” or “ys of age” or “yrs of age” or “y of age” or “year? old” or “yr old” or “ys old” or “yrs old” or “y old”)) or child* or kid or kids or girl or girls or girlhood or boy or boys or boyhood or juvenil* or prepube* or pubert* or pubescent? or pubescence or preadolesc* or youth? or (young adj2 (people* or person* or individual* or population*)) or schoolage? or “school age?” or schoolchild* or “school child*” or “1st grade*” or “first grade*” or “grade? 1” or “grade? one” or “2nd grade*” or “second grade*” or “grade? 2” or “grade? two” or “3rd grade*” or “third grade*” or “grade? 3” or “grade? three” or “4th grade*” or “fourth grade*” or “grade? 4” or “grade? four” or “5th grade*” or “fifth grade*” or “grade? 5” or “grade? five” or “6th grade*” or “sixth grade*” or “grade? 6” or “grade? six” or “7th grade*” or “seventh grade*” or “grade? 7” or “grade? seven” or school* or “pre school*” or “elementary school*” or “primary school*” or “postprimary school*” or “grade school*” or “middle school*” or “junior high” or “intermedia* school*” or “presecondary school*” or preschool* or gradeschool* or middleschool* or kin#ergar#en* or “elementary education*” or “primary education*” or “postprimary education*” or “intermedia* education*” or “presecondary education*” or “secondary education*” or “k through 12” or “k through twelve” or “k thru 12” or “k thru twelve” or “k 12” or k12 or “k twelve” or p?ediat*).ti,ab,kf.
2. exp preventive health service/ or preventive medicine/ or pc.fs. or (checkup? or “check up?” or “well check?” or “wellcheck?” or ((“well child” or “health supervision” or “primary care”) and visit*) or (preven??tive adj (medicine or health or care or service? or program?)) or “health education” or “health promotion”).ti,ab,kf.
3. clinical practice/ or doctor patient relationship/ or clinical competence/ or (“practice pattern?” or “prescri* pattern?” or “patient relation*” or “physician patient” or “patient physician” or “doctor patient” or “patient doctor” or content or ((physician? or doctor? or clinician?) adj behavior?)).ti,ab,kf. or ((exp Communication/ or communicat*.ti,ab,kf.) and (Patients/ or (patient? or inpatient? or outpatient? or client?).ti,ab,kf.))
4. general practitioner/ or exp primary health care/ or (p?ediatrician? or (p?ediatric? adj (clinic? or practice? or setting? or hospital? or physician? or doctor? or clinician? or practice? or health))).ti,ab,kf,hw,in. or (children?? adj3 hospital?).ti,ab,kf,in. or ((“primary care” adj (provider? or physician? or doctor? or clinician? or clinic?)) or (family adj (physician? or doctor? or clinician? or clinic?))).ti,ab,kf.
5. ((exp animal/ or exp plant/ or nonhuman/ or exp animal experiment/ or exp experimental organism/ or exp animal model/) not ((exp animal/ or exp plant/ or nonhuman/ or exp animal experiment/ or exp experimental organism/ or exp animal model/) and (exp human/ or exp human experiment/))) or (((animal? or nonhuman? or “non human?”) adj2 (study or studies or model* or experiment*)) or mouse or mice or rat or rats or monkey or monkeys or “preclinical study” or “preclinical studies” or “pre clinical study” or “pre clinical studies”).ti.
6. (1 and 2 and 3 and 4) not 5
7. limit 6 to (english language and yr=“2014-Current”)

### CENTRAL (Cochrane Central Register of Controlled Trials) Search

#### Search run August 5, 2025

#1 [mh Child] OR [mh ^Puberty] OR ((age?:ti,ab,kw NEAR/2 (6:ti,ab,kw OR 7:ti,ab,kw OR 8:ti,ab,kw OR 9:ti,ab,kw OR 10:ti,ab,kw OR 11:ti,ab,kw OR 12:ti,ab,kw OR six:ti,ab,kw OR seven:ti,ab,kw OR eight:ti,ab,kw OR nine:ti,ab,kw OR ten:ti,ab,kw OR eleven:ti,ab,kw OR twelve:ti,ab,kw)) OR ((6:ti,ab,kw OR 7:ti,ab,kw OR 8:ti,ab,kw OR 9:ti,ab,kw OR 10:ti,ab,kw OR 11:ti,ab,kw OR 12:ti,ab,kw OR six:ti,ab,kw OR seven:ti,ab,kw OR eight:ti,ab,kw OR nine:ti,ab,kw OR ten:ti,ab,kw OR eleven:ti,ab,kw OR twelve:ti,ab,kw) NEAR/2 ((year? NEXT “of age”):ti,ab,kw OR “yr of age”:ti,ab,kw OR “ys of age”:ti,ab,kw OR “yrs of age”:ti,ab,kw OR “y of age”:ti,ab,kw OR (year? NEXT “old”):ti,ab,kw OR “yr old”:ti,ab,kw OR “ys old”:ti,ab,kw OR “yrs old”:ti,ab,kw OR “y old”:ti,ab,kw)) OR child*:ti,ab,kw OR kid:ti,ab,kw OR kids:ti,ab,kw OR girl:ti,ab,kw OR girls:ti,ab,kw OR girlhood:ti,ab,kw OR boy:ti,ab,kw OR boys:ti,ab,kw OR boyhood:ti,ab,kw OR juvenil*:ti,ab,kw OR prepube*:ti,ab,kw OR pubert*:ti,ab,kw OR pubescent?:ti,ab,kw OR pubescence:ti,ab,kw OR preadolesc*:ti,ab,kw OR youth?:ti,ab,kw OR (young:ti,ab,kw NEAR/2 (people*:ti,ab,kw OR person*:ti,ab,kw OR individual*:ti,ab,kw OR population*:ti,ab,kw)) OR schoolage?:ti,ab,kw OR (“school” NEXT age?):ti,ab,kw OR schoolchild*:ti,ab,kw OR (“school” NEXT child*):ti,ab,kw OR (“1st” NEXT grade*):ti,ab,kw OR (“first” NEXT grade*):ti,ab,kw OR (grade? NEXT “1”):ti,ab,kw OR (grade? NEXT “one”):ti,ab,kw OR (“2nd” NEXT grade*):ti,ab,kw OR (“second” NEXT grade*):ti,ab,kw OR (grade? NEXT “2”):ti,ab,kw OR (grade? NEXT “two”):ti,ab,kw OR (“3rd” NEXT grade*):ti,ab,kw OR (“third” NEXT grade*):ti,ab,kw OR (grade? NEXT “3”):ti,ab,kw OR (grade? NEXT “three”):ti,ab,kw OR (“4th” NEXT grade*):ti,ab,kw OR (“fourth” NEXT grade*):ti,ab,kw OR (grade? NEXT “4”):ti,ab,kw OR (grade? NEXT “four”):ti,ab,kw OR (“5th” NEXT grade*):ti,ab,kw OR (“fifth” NEXT grade*):ti,ab,kw OR (grade? NEXT “5”):ti,ab,kw OR (grade? NEXT “five”):ti,ab,kw OR (“6th” NEXT grade*):ti,ab,kw OR (“sixth” NEXT grade*):ti,ab,kw OR (grade? NEXT “6”):ti,ab,kw OR (grade? NEXT “six”):ti,ab,kw OR (“7th” NEXT grade*):ti,ab,kw OR (“seventh” NEXT grade*):ti,ab,kw OR (grade? NEXT “7”):ti,ab,kw OR (grade? NEXT “seven”):ti,ab,kw OR school*:ti,ab,kw OR (“pre” NEXT school*):ti,ab,kw OR (“elementary” NEXT school*):ti,ab,kw OR (“primary” NEXT school*):ti,ab,kw OR (“postprimary” NEXT school*):ti,ab,kw OR (“grade” NEXT school*):ti,ab,kw OR (“middle” NEXT school*):ti,ab,kw OR “junior high”:ti,ab,kw OR (intermedia* NEXT school*):ti,ab,kw OR (“presecondary” NEXT school*):ti,ab,kw OR preschool*:ti,ab,kw OR gradeschool*:ti,ab,kw OR middleschool*:ti,ab,kw OR kin?ergar?en*:ti,ab,kw OR (“elementary” NEXT education*):ti,ab,kw OR (“primary” NEXT education*):ti,ab,kw OR (“postprimary” NEXT education*):ti,ab,kw OR (intermedia* NEXT education*):ti,ab,kw OR (“presecondary” NEXT education*):ti,ab,kw OR (“secondary” NEXT education*):ti,ab,kw OR “k through 12”:ti,ab,kw OR “k through twelve”:ti,ab,kw OR “k thru 12”:ti,ab,kw OR “k thru twelve”:ti,ab,kw OR “k 12”:ti,ab,kw OR k12:ti,ab,kw OR “k twelve”:ti,ab,kw OR p?ediat*:ti,ab,kw)
#2 [mh “Preventive Health Services”] OR [mh ^”Preventive Medicine”] OR [mh /PC] OR (checkup?:ti,ab,kw OR (“check” NEXT up?):ti,ab,kw OR (“well” NEXT check?):ti,ab,kw OR wellcheck?:ti,ab,kw OR ((“well child”:ti,ab,kw OR “health supervision”:ti,ab,kw OR “primary care”:ti,ab,kw) AND visit*:ti,ab,kw) OR (preven??tive:ti,ab,kw NEXT (medicine:ti,ab,kw OR health:ti,ab,kw OR care:ti,ab,kw OR service?:ti,ab,kw OR program?:ti,ab,kw)) OR “health education”:ti,ab,kw OR “health promotion”:ti,ab,kw)
#3 [mh ^”Practice Patterns, Physicians’”] OR [mh ^”Physician-Patient Relations”] OR [mh ^”Clinical Competence”] OR ((“practice” NEXT pattern?):ti,ab,kw OR (prescri* NEXT pattern?):ti,ab,kw OR (“patient” NEXT relation*):ti,ab,kw OR “physician patient”:ti,ab,kw OR “patient physician”:ti,ab,kw OR “doctor patient”:ti,ab,kw OR “patient doctor”:ti,ab,kw OR content:ti,ab,kw OR ((physician?:ti,ab,kw OR doctor?:ti,ab,kw OR clinician?:ti,ab,kw) NEXT behavior?:ti,ab,kw)) OR (([mh Communication] OR communicat*:ti,ab,kw) AND ([mh ^Patients] OR (patient?:ti,ab,kw OR inpatient?:ti,ab,kw OR outpatient?:ti,ab,kw OR client?:ti,ab,kw)))
#4 [mh ^”Physicians, Primary Care”] OR [mh ^”Primary Health Care”] OR (p?ediatrician?:ti,ab,kw OR (p?ediatric?:ti,ab,kw NEXT (clinic?:ti,ab,kw OR practice?:ti,ab,kw OR setting?:ti,ab,kw OR hospital?:ti,ab,kw OR physician?:ti,ab,kw OR doctor?:ti,ab,kw OR clinician?:ti,ab,kw OR practice?:ti,ab,kw OR health:ti,ab,kw))) OR (children??:ti,ab,kw NEAR/3 hospital?:ti,ab,kw) OR ((“primary care”:ti,ab,kw NEXT (provider?:ti,ab,kw OR physician?:ti,ab,kw OR doctor?:ti,ab,kw OR clinician?:ti,ab,kw OR clinic?:ti,ab,kw)) OR (family:ti,ab,kw NEXT (physician?:ti,ab,kw OR doctor?:ti,ab,kw OR clinician?:ti,ab,kw OR clinic?:ti,ab,kw)))
#5 ([mh Animals] NOT ([mh Animals] AND [mh ^Humans])) OR [mh “Animal Experimentation”] OR [mh “Models, Animal”] OR ((animal?:ti NEAR/2 (study:ti OR model*:ti OR experiment*:ti)) OR mouse:ti OR mice:ti OR rat:ti OR rats:ti OR monkey:ti OR monkeys:ti OR “preclinical study”:ti)
#6 (#1 AND #2 AND #3 AND #4) NOT #5 with Publication Year from 2014 to 2025, in Trials

## Additional file 2. Data extraction template

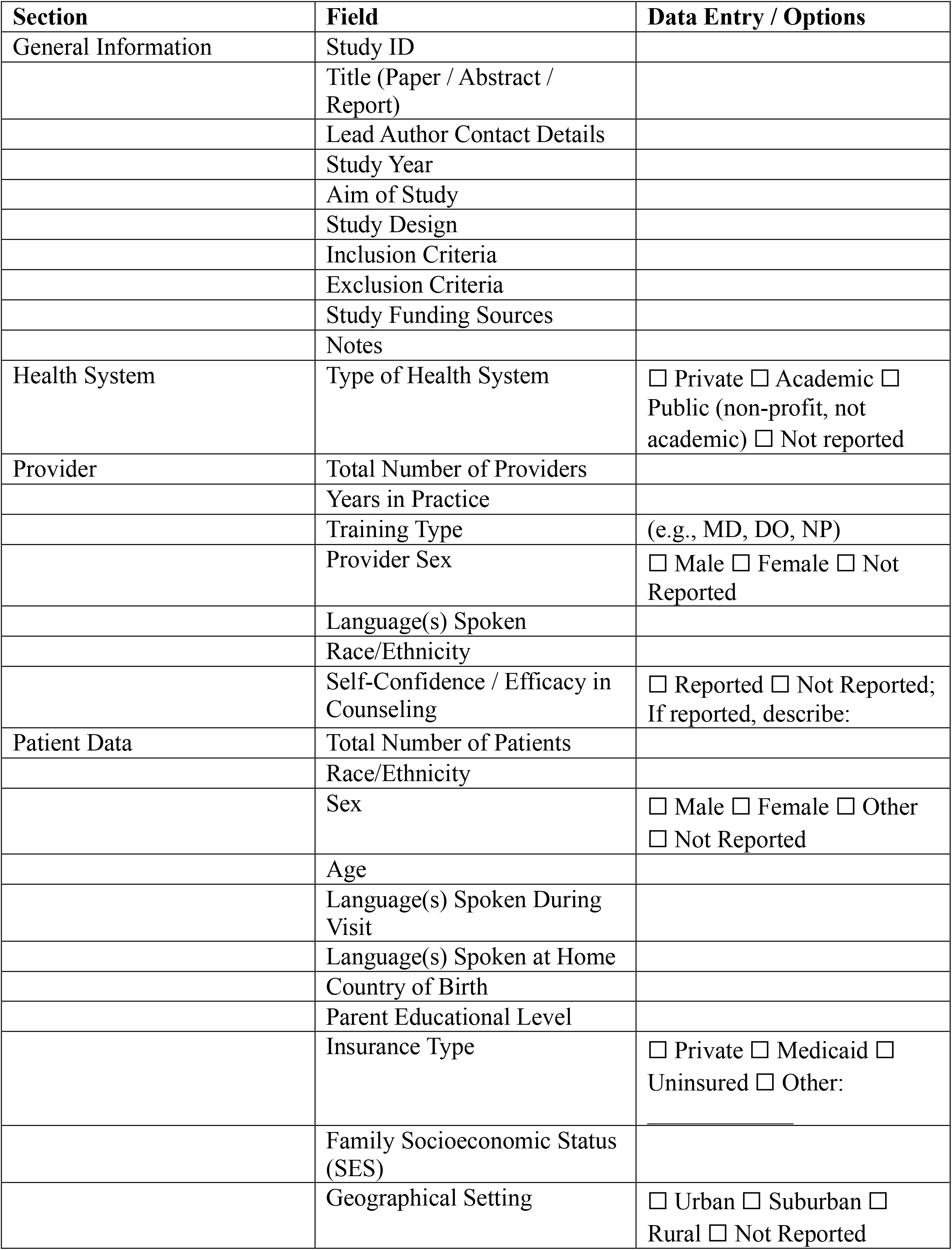
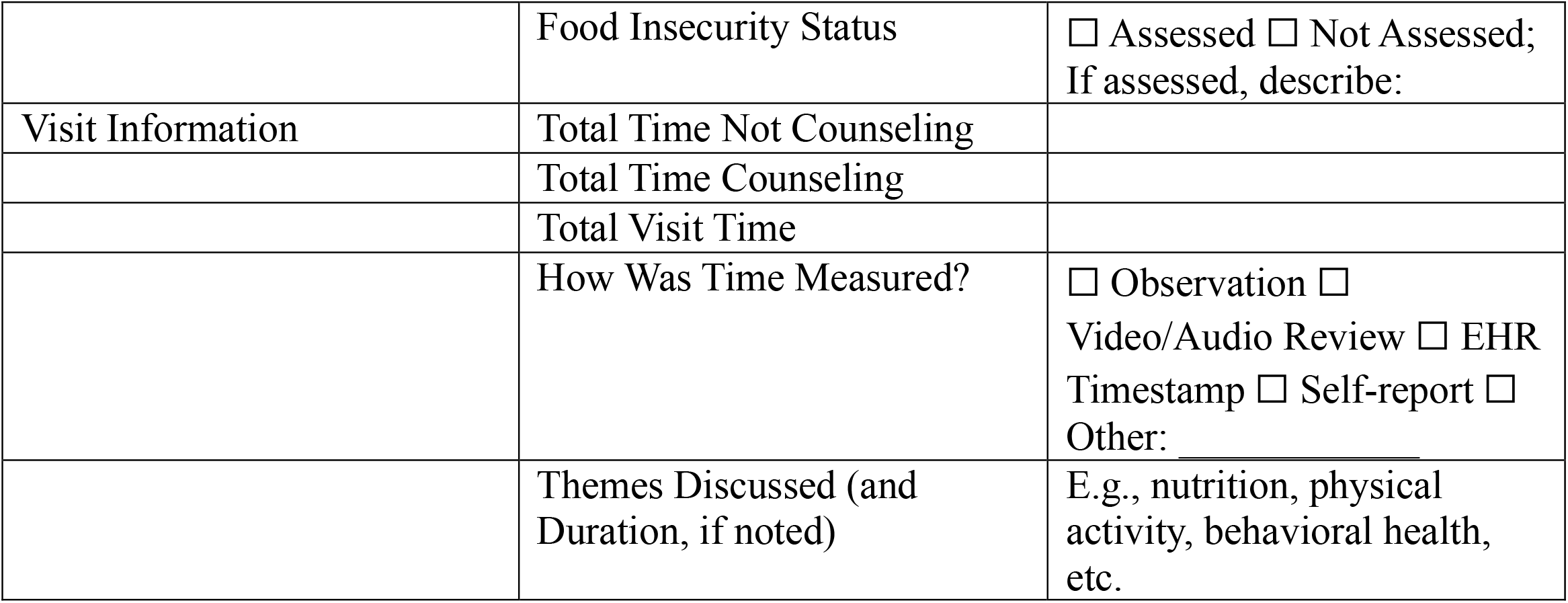

| Section | Field | Data Entry / Options |
| --- | --- | --- |
| General Information | Study ID |  |
|  | Title (Paper / Abstract / Report) |  |
|  | Lead Author Contact Details |  |
|  | Study Year |  |
|  | Aim of Study |  |
|  | Study Design |  |
|  | Inclusion Criteria |  |
|  | Exclusion Criteria |  |
|  | Study Funding Sources |  |
|  | Notes |  |
| Health System | Type of Health System | ☐ Private ☐ Academic ☐ Public (non-profit, not academic) ☐ Not reported |
| Provider | Total Number of Providers |  |
|  | Years in Practice |  |
|  | Training Type | (e.g., MD, DO, NP) |
|  | Provider Sex | ☐ Male ☐ Female ☐ Not Reported |
|  | Language(s) Spoken |  |
|  | Race/Ethnicity |  |
|  | Self-Confidence / Efficacy in Counseling | ☐ Reported ☐ Not Reported; If reported, describe: |
| Patient Data | Total Number of Patients |  |
|  | Race/Ethnicity |  |
|  | Sex | ☐ Male ☐ Female ☐ Other ☐ Not Reported |
|  | Age |  |
|  | Language(s) Spoken During Visit |  |
|  | Language(s) Spoken at Home |  |
|  | Country of Birth |  |
|  | Parent Educational Level |  |
|  | Insurance Type | ☐ Private ☐ Medicaid ☐ Uninsured ☐ Other: |
|  | Family Socioeconomic Status (SES) |  |
|  | Geographical Setting | ☐ Urban ☐ Suburban ☐ Rural ☐ Not Reported |
|  | Food Insecurity Status | ☐ Assessed ☐ Not Assessed;<br>If assessed, describe: |
| Visit Information | Total Time Not Counseling |  |
|  | Total Time Counseling |  |
|  | Total Visit Time |  |
|  | How Was Time Measured? | ☐ Observation ☐<br>Video/Audio Review ☐ EHR<br>Timestamp ☐ Self-report ☐<br>Other: _____ |
|  | Themes Discussed (and<br>Duration, if noted) | E.g., nutrition, physical<br>activity, behavioral health,<br>etc. |

